# Measuring elimination of *gambiense* human African trypanosomiasis: A comparison of deceptively different metrics

**DOI:** 10.1101/2025.08.11.25331730

**Authors:** Samuel A. Sutherland, Jason J. Madan, Kat S. Rock

**Affiliations:** The Zeeman Institute for Systems Biology & Infectious Disease Epidemiology Research, University of Warwick, University Road, Coventry, CV4 7AL, United Kingdom; Centre for Health Economics at Warwick, Warwick Medical School, University of Warwick, University Road, Coventry, CV4 7AL, United Kingdom; Warwick Mathematics Institute, University of Warwick, University Road, Coventry, CV4 7AL, United Kingdom

**Keywords:** elimination, human African trypanosomiasis (HAT), sleeping sickness, modelling, neglected tropical diseases (NTDs), disease monitoring, disease surveillance, terminology, World Health Organisation (WHO)

## Abstract

Modelling is an effective and widely used tool for predicting the future trajectory of infectious diseases. One of its strengths is its ability to measure and predict metrics that are unobservable in real life, such as the true prevalence, or when infection events occur. Policy goals, on the other hand, must necessarily be based on observable metrics in order to be verifiable. The World Health Organisation (WHO) has targeted *gambiense* human African trypanosomiasis (gHAT) for elimination of transmission (EoT) by 2030. In order to verify this, the WHO HAT Elimination Technical Advisory Group (HAT-e-TAG) have recently defined the country-level indicator for EoT as five years of no new locally-infected reported cases, along with a sufficient level of surveillance. While this indicator is a useful and concrete metric that can be clearly measured, it does not directly measure the actual transmission or prevalence of the disease. In some cases, the timing of the last transmission event or the point when infection is no longer present in a country may differ quite substantially from when the country reaches the WHO indicator for elimination of transmission. In this article, we discuss the difference between these different metrics and show with some examples how much they can disagree. Furthermore, modelling papers have previously been published using various approximations of EoT, which has the potential to cause confusion. In this paper, we highlight the potential misunderstandings which could occur due to seemingly trivial differences in the definition of elimination and its indicators, especially in the context of such a slow-progressing disease as gHAT. We conclude that while there is value in models predicting both observable and unobservable metrics, modellers need to ensure that they are clear about definitions when communicating results, and policymakers need to be sure that they know what definitions are being used when drawing conclusions from model results. While this paper focuses specifically on three indicators of EoT for gHAT, the general points made here could have applications in other infections approaching the endgame, such as onchocerciasis, or even beyond public health.

## Defining elimination of gHAT

*Gambiense* human African trypanosomiasis (*g*HAT) is a vector-borne parasitic infection endemic in areas of West and Central Africa. If it is not treated for several years, infection usually, although not always, leads to death [1]. There is no vaccine to protect against infection and infection control efforts usually rely on detection and treatment, sometimes with vector control to reduce transmission opportunities from tsetse vectors. *g*HAT was targeted by the World Health Organization (WHO) for elimination as a public health problem (EPHP) by 2020 [2]—a goal which narrowly missed its target year [3] but was attained just two years later [4]—and has also been marked for elimination of transmission (EoT) to humans by 2030 as part of the *road map for neglected tropical diseases (NTDs) 2021–2030* [5]. Whilst the WHO indicator for elimination as a public health problem (WHO-EPHP) is completely defined by measurable case reporting, EoT is unfortunately trickier to measure. It is not possible to directly observe the transmission events causing new infections, so a proxy indicator is needed to be able to measure progress towards this target. In particular, *g*HAT has a very long infection time (often multiple years, sometimes over a decade [6]) meaning that cases are often found substantially after the patient was initially infected.

In November 2023, the WHO‘s HAT elimination Technical Advisory Group (HAT-e-TAG) published its new set of criteria which will be used to assess whether countries have met the threshold for verification of EoT [7]. EoT indicators are different for different infections and depend on various factors including duration of infection, diagnostic methods available to detect the pathogen and feasible monitoring strategies [5]. E.g. the country-level EoT indicator for onchocerciasis includes measuring prevalence in the black fly vector as well as sampling in children—specifically, *<*0.1% Immunoglobulin G4 (IgG4) antibody seropositivity to the Ov-16 *Onchocerca volvulus* antigen in children younger than 10 years, and *<*0.05% positivity by pool screen Polymerase Chain Reaction (PCR) in at least 6,000 wild-caught flies (heads only)[8]. For *g*HAT, the indicator defined by the HAT-e-TAG for EoT in a given country is five years of zero reports of new locally-infected cases in the country, subject to sufficient case-finding activities as specified in their full guidelines [7].

Cases of *g*HAT are typically identified by an initial serological screening test, either the card agglutination test for trypanosomiasis (CATT) or a rapid diagnostic test (RDT), followed by confirmatory microscopy to visualise the parasite in the blood. All currently available treatments may have substantial side effects, so treatment is only able to be given to patients with confirmed infection by the parasite meaning mass drug administration (MDA) is not presently an option. While treatment was improved in 2020 for most cases with the rollout of the oral drug fexinidazole, this is still generally only indicated for use in confirmed cases [10], although some countries will treat strong serological suspects. Additionally, some cases still need to receive intravenous medication in the hospital depending on age, weight or in the case of severe disease [10]. More recently, a new medication known as acoziborole has been developed [11] and trials are currently ongoing to evaluate its safety for use in suspected cases [12, 13]. Even so, acoziborole is not currently planned for use in patients without at least serological suspicion of *g*HAT.

Cases are typically diagnosed through one of two routes – either “active” or “passive” screening. Active screening consists of mass screening of willing participants in at-risk areas, either by CATT or RDTs [14]. Passive screening refers to infected people presenting themselves at fixed health facilities with specific symptoms of *g*HAT disease, and subsequently being tested for it [15]. This relies on both the availability of RDTs at the facility where the patient presents and the sensitisation of the health care provider to *g*HAT symptoms. This means that detection is necessarily highly linked to intervention, in contrast to many other NTDs where MDA is the primary intervention and surveillance generally occurs as a separate process.

A previous article described some of the challenges associated with measuring EoT for *g*HAT using case reporting [16], stressing that the level of active and passive case finding activities and presence or absence of vector control could influence EoT measurement; however, at the time of publication, the formal WHO indicator for *g*HAT had not been published.

In this report, we aim to better understand how the WHO-EoT indicator (which can be directly measured), is linked to LTE and NRI (as defined in Table 1) which cannot be directly observed. We evaluate this using a modelling framework, comparing the point at which the WHO indicator is attained, the point at which transmission truly ceases, and the point at which no infection remains. We also aim to clarify the difference between the WHO-EoT indicator and the definition of EoT used in previous modelling papers by our group and other authors.

**Table 1.**
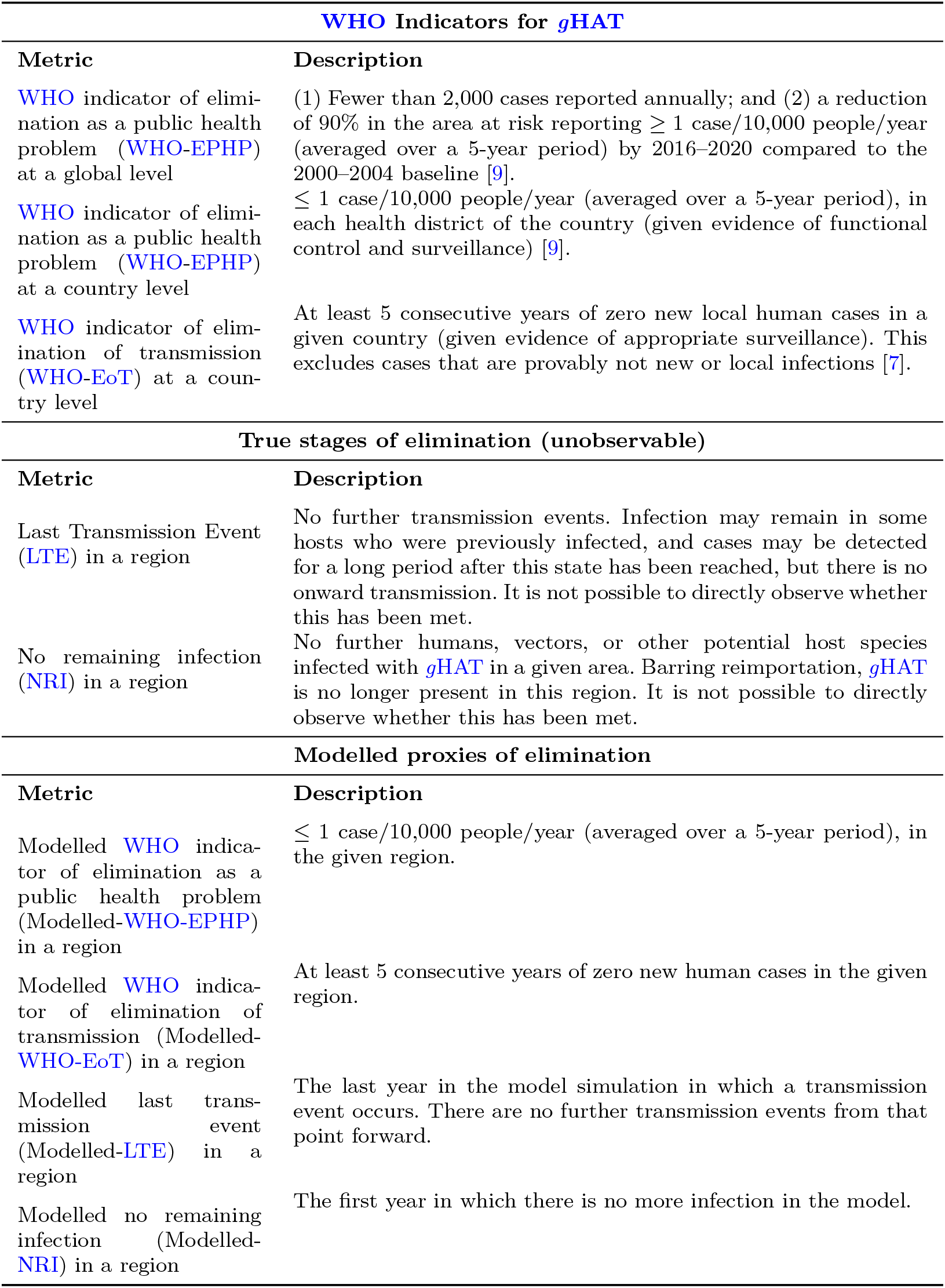
List of *g*HAT elimination metrics and their definitions.

| WHO Indicators for *gHAT* |  |
| --- | --- |
| Metric | Description |
| WHO indicator of elimination as a public health problem (WHO-EPHP) at a global level | (1) Fewer than 2,000 cases reported annually; and (2) a reduction of 90% in the area at risk reporting $\geq 1$ case/10,000 people/year (averaged over a 5-year period) by 2016–2020 compared to the 2000–2004 baseline [9]. |
| WHO indicator of elimination as a public health problem (WHO-EPHP) at a country level | $\leq 1$ case/10,000 people/year (averaged over a 5-year period), in each health district of the country (given evidence of functional control and surveillance) [9]. |
| WHO indicator of elimination of transmission (WHO-EoT) at a country level | At least 5 consecutive years of zero new local human cases in a given country (given evidence of appropriate surveillance). This excludes cases that are provably not new or local infections [7]. |
| True stages of elimination (unobservable) |  |
| Metric | Description |
| Last Transmission Event (LTE) in a region | No further transmission events. Infection may remain in some hosts who were previously infected, and cases may be detected for a long period after this state has been reached, but there is no onward transmission. It is not possible to directly observe whether this has been met. |
| No remaining infection (NRI) in a region | No further humans, vectors, or other potential host species infected with *gHAT* in a given area. Barring reimportation, *gHAT* is no longer present in this region. It is not possible to directly observe whether this has been met. |
| Modelled proxies of elimination |  |
| Metric | Description |
| Modelled WHO indicator of elimination as a public health problem (Modelled-WHO-EPHP) in a region | $\leq 1$ case/10,000 people/year (averaged over a 5-year period), in the given region. |
| Modelled WHO indicator of elimination of transmission (Modelled-WHO-EoT) in a region | At least 5 consecutive years of zero new human cases in the given region. |
| Modelled last transmission event (Modelled-LTE) in a region | The last year in the model simulation in which a transmission event occurs. There are no further transmission events from that point forward. |
| Modelled no remaining infection (Modelled-NRI) in a region | The first year in which there is no more infection in the model. |

In this article, we will be focusing specifically on *g*HAT, however, the general points made here are applicable to all infectious diseases targeted for EoT or eradication.

### Some illustrative examples

Figure 1 shows two example potential infection histories for a hypothetical location approaching *g*HAT elimination. In *Example 1*, we see the progression of transmission and infection in an area with effective active and passive screening and low levels of transmission, potentially due to vector control. Here, we can see that the last transmission occurs in 2025, however, this infected person is only detected around two years later in early 2027. Applying the rule of the WHO indicator for EoT, this would mean that the first of the five years of no case reporting could not start until 2028, finally attaining the five years of no cases for WHO-EoT in 2033, five years after there is no latent infection, and seven years after the last transmission event.

**Fig. 1.**
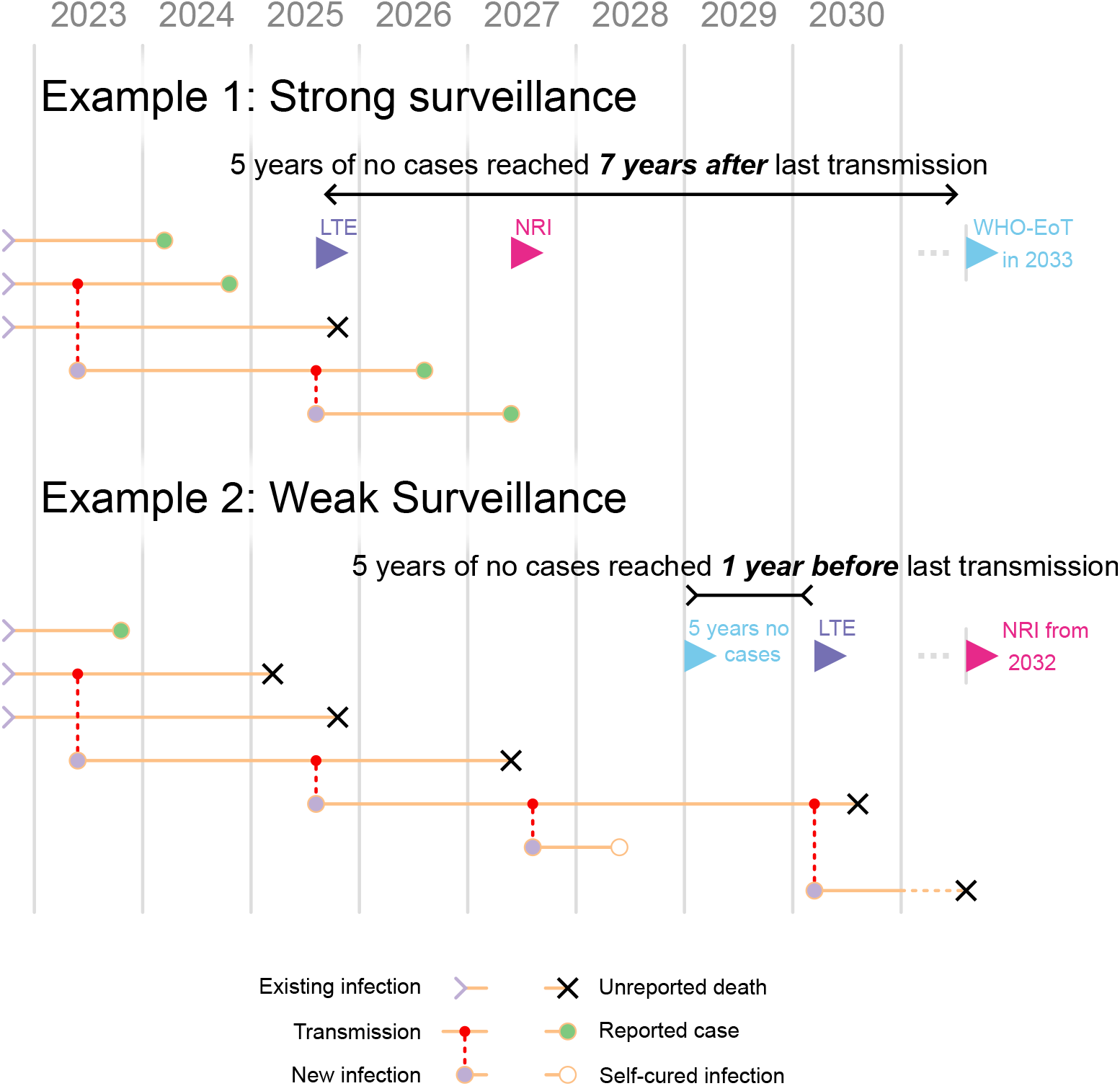
Two illustrations of possible pathways to country-level elimination starting from very low prevalence of *g*HAT infection. All examples start with three extant infections in 2022 and the corresponding horizontal orange lines show individuals continuing to be infectious. We assume that infection ends in either detection and subsequent treatment (marked by green circles), an unreported death, (marked by black crosses), or a potential self-curing infection which is not reported (marked by an unfilled circle). Coloured flags mark the achievement of the three different elimination metrics: five years of no cases/WHO-EoT (blue), LTE (purple), and NRI (pink). Note that in the second example we refer to “5 years no cases” instead of WHO-EoT because it is unlikely that such a setting would meet the criteria for the WHO metric of EoT due to insufficient surveillance.

In *Example 2*, we see an example of the same area with weaker interventions, consisting of less effective screening, where there is still ongoing transmission more than five years after the last reported case. Because of this, LTE actually occurs after five years of no reported cases, and after the LTE, there is still a remaining infected person for another couple of years. Since the WHO indicator is based on case reporting, it is strongly dependent on the level of surveillance. This highlights the importance of the surveillance component of the definition of WHO-EoT, since without sufficient monitoring, it is impossible to tell the difference between there being no infection and all the infection being missed.

### Modelling elimination and its indicators

Mathematical modelling has been used to address a range of quantitative questions about *g*HAT transmission including assessing the success of historical interventions on transmission and case reporting, evaluating the expected impact of a range of future strategies on infection dynamics, and analysing the costs and cost-effectiveness of intervention strategies. While metrics like LTE and EoI are not directly measurable in the real world, we can use model simulations to compute these metrics from the model outputs. Similarly, we can estimate the associated time until WHO-EoT using the simulated case reporting.

Our modelling team have previously developed and published a model of *g*HAT transmission which we have fitted to case data and used in a variety of settings to simulate infection dynamics [17–22]. Our model is a compartmental transmission model with explicit modelling of human and tsetse dynamics, as well as risk stratification in humans and the potential for non-human animal or human asymptomatic hosts. In this paper, we use a stochastic version of our model that incorporates chance events and outputs integer numbers of infections and cases. A stochastic model was chosen here since the integer numbers of people in each infection state and event means that both Modelled-LTE and Modelled-EoI can be directly read from when the appropriate values reach zero in the model. Previous work has compared the effectiveness of different interventions in various settings using variants of this model. Work by the authors of this paper has previously used a deterministic model, which is simpler to simulate and reflects average dynamics, however this type of model cannot reach exactly zero infections and so needs a proxy threshold to determine when elimination is reached. This technique generally gives similar results for expected year of EoT, however a stochastic model like that used in this paper generally produces a broader range of predictions for EoT, i.e. more uncertainty [23].

Among the outputs of the model are estimates of the probability of reaching different indicators of EoT by each given year. For this present work, we have used the model to output each of the region-specific metrics detailed in Table 1, using projections based on an exemplar region, which still reflects “typical” *g*HAT transmission. For simplicity, we have not included the possibility of asymptomatic human infections in these projections, although doing so would not change the illustrative points we will highlight. We have included the possibility of animal infections in an ensemble model as in [24].

In order to evaluate and compare the different metrics of elimination, we ran simulations under different intervention scenarios and recorded when each metric of elimination was reached in each realisation. We considered eight different hypothetical settings with a fixed population size of 100,000 starting from the endemic equilibrium in year 0, and with parameters based on our existing model fits for one of eight different health zones of the Democratic Republic of the Congo (DRC) to capture a range of plausible infection dynamics and to confirm that our results would be qualitatively similar even if parameters such as the tsetse-to-human ratio or the time to passive detection were varied. We ran these simulations for two years under a weak level of passive screening only, then simulated from year 2 to year 200 under three different strategies, covering different levels of passive screening, active screening, and vector control; the first has 10% of the population screened in active screening each year, vector control with a 80% reduction after one year and effective passive screening (comparable to present-day coverage, with an average time from infection to detection of 2.7 years if not found in active screening under the parameterisation used in figure 2), the second is the same, but with no vector control, and the third has worse surveillance with only 2% active screening coverage per year and weak passive screening (comparable to coverage estimated for the early 2000s, the average time from infection to detection of 3.3 years if not found in active screening under the parameterisation used in figure 2). As we have not directly used case trends or intervention history matched to any real locations, this illustrative modelling analysis does not reflect progress or predictions for any specific health zones in the DRC.

**Fig. 2.**
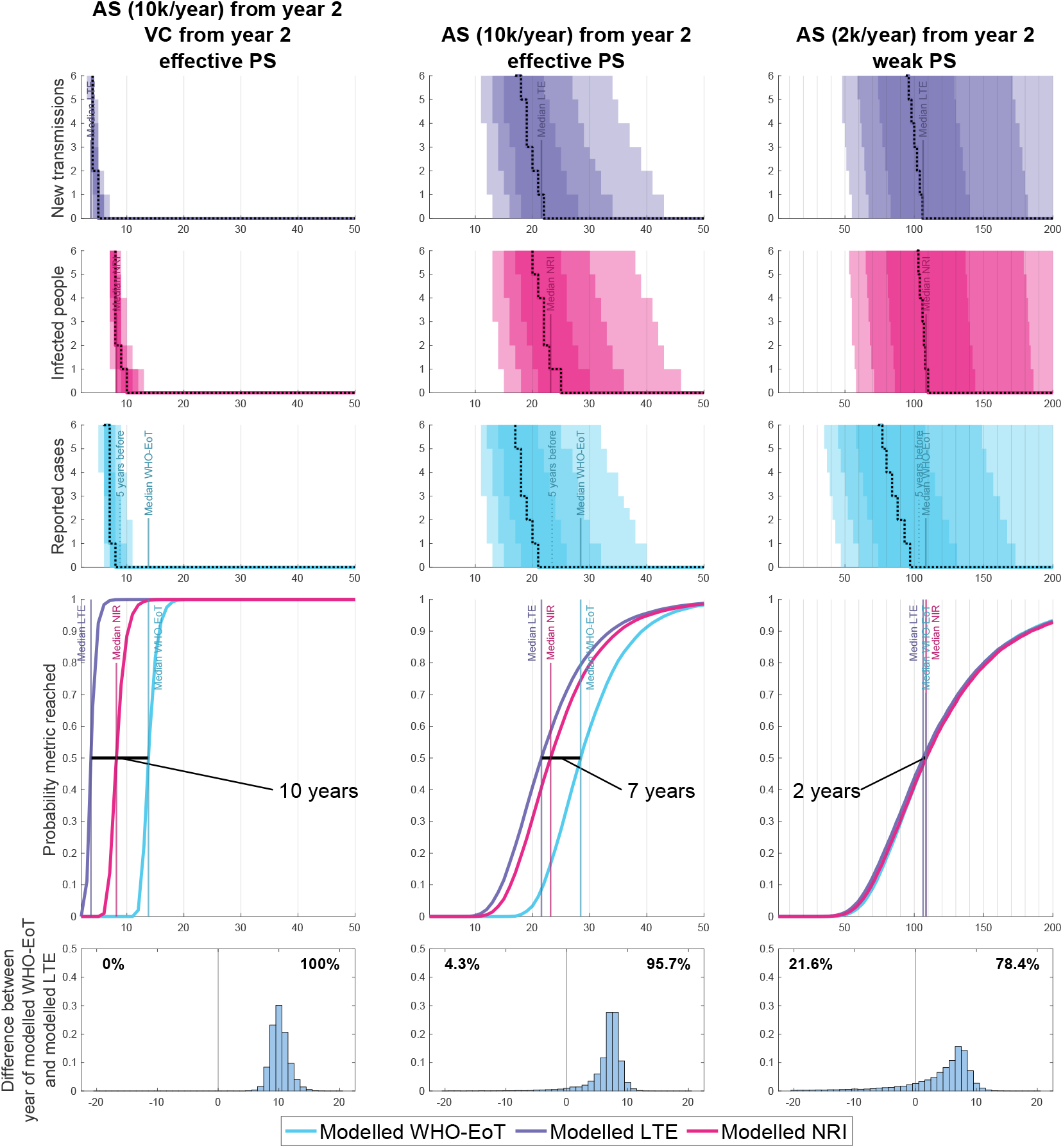
Model simulations showing various outputs and how they link to three different elimination metrics. The top three figures in each column show the number of new infections (purple) and the number of cases each year (blue), as well as the number of infections remaining at any given time (pink). The solid lines show the median model output, with 50%, 80%, and 95% prediction intervals shown by the shaded regions. The fourth figure shows estimates of the probability of Modelled-WHO-EoT, Modelled-LTE and Modelled-NRI by year, which are computed from the proportion of the 20,000 stochastic model simulations which have achieved the specific metric. The bottom figure shows a histogram of the delay between Modelled-LTE and WHO-EoT. The percentages show the proportion of realisations in which the final transmission event comes respectively after or before the five years of no cases required for WHO-EoT. The parameters used to generate the simulations used in this figure were based on our model parameterisation for Bagata health zone in Bandundu Nord coordination in the DRC, although since we have not directly used case trends or intervention history here, this analysis does not reflect progress or predictions for the real health zone.

Figure 2 shows the results in one setting for the strategies ranging from heavy intervention to minimal intervention. We can see in the figure that across the different strategies, the Modelled-WHO-EoT metric is conservative compared to Modelled-LTE and Modelled-NRI. The exact size of the delay depends on the location-specific infection drivers and the level of intervention, but across the majority of our simulations, the order is the same. Firstly, Modelled-LTE is reached, followed by Modelled-NRI, finally followed by Modelled-WHO-EoT. This is because NRI must necessarily come after LTE, and in most of the scenarios with more effective surveillance, the final infection is more likely to be reported as a case than to die unreported without causing another infection. We also see that in all the scenarios with a good level of passive screening, Modelled-LTE is almost certain to occur before Modelled-WHO-EoT, with an average delay of around 7 years without vector control, and 10 years with with vector control. Results for the seven other settings are found in our Supplementary Information and show qualitatively similar behaviour. If passive surveillance is weak, then there is also a moderate probability that there could be 5 years of no case reporting before the LTE, which is why we consider effective surveillance an important stipulation of the WHO-EoT indicator.

### Cross-discipline language barriers and interpretation for policy

Our modelling has shown that these different metrics, despite all nominally being elimination metrics of some kind, can be reached at quite different times. Policymakers should be very aware of how modellers are simulating EoT in their outputs if they want to use the results—are they using the WHO-EoT indicator definition for EoT or are they modelling LTE or NRI? For example, one modelling study for onchocerciasis specifies that they are using a definition closest to what is referred to in this present report as NRI, specifically an “absence of parasites in humans and flies 50 years after treatment cessation”. Studies including predictions of *g*HAT elimination by the “HAT Modelling and Economic Predictions for Policy (HAT MEPP)“ group, with which the authors of this paper are affiliated, have previously defined EoT as the first year after “the last simulated transmission event to humans”, equivalent here to one year after the Modelled-LTE (recent examples include [19–22]), which is likely not to align with when the WHO-EoT *g*HAT indicator will be met. In the future the HAT MEPP group will transition to reporting the Modelled-LTE and Modelled-WHO-EoT to make the distinction more clear between predictions for the transmission events and case reporting.

Broadly speaking the NTD community need to be careful to not conflate the same words to mean slightly different things—e.g. “elimination” (with no qualifier) could be EPHP or EoT—although in recent years this has improved with the WHO leading on producing clearer definitions and updating older documentation. Modellers should also do more to bridge the language gap between describing modelling outputs and how clinicians or policymakers understand the terminology; the recent publication of the WHO-EoT indicator for *g*HAT [7] has now made the distinction between these different metrics far easier to explain. We would recommend that modellers producing projections of elimination should be very clear what definitions they are using, or even publish projections using multiple indicators. Furthermore, modellers may also talk about local or regional EoT, whereas the WHO indicators are currently only defined at the country level, so attention should be taken to the spatial scale being considered.

While all these metrics have merit as measures of progress, it is vital that policymakers, modellers, and other stakeholders alike are clear on what metric they are using and that false equivalencies are not drawn between two metrics that may initially appear similar but are not, in fact, the same.

## Supporting information

Supplementary results

## Data Availability

Full source code, data used and full results are available at http://doi.org/10.17605/OSF.IO/N4DKZ

## List of Abbreviations

*g*HAT: *gambiense* human African trypanosomiasis
CATT: card agglutination test for trypanosomiasis
DRC: Democratic Republic of the Congo
EoT: elimination of transmission
EPHP: elimination as a public health problem
HAT MEPP: HAT Modelling and Economic Predictions for Policy
HAT-e-TAG: HAT elimination Technical Advisory Group
IgG4: Immunoglobulin G4
LTE: last transmission event
MDA: mass drug administration
NRI: no remaining infection
NTD: neglected tropical disease
PCR: Polymerase Chain Reaction
RDT: rapid diagnostic test
WHO: World Health Organization

## Declarations

### Ethics approval and consent to participate

This simulation study did not directly use any human case data and used previously published model parameterisation from other publicly available modelling articles. No new data collection took place within the scope of this modelling study.

### Consent for publication

Not applicable

### Availability of data and materials

Full source code, data used and full results are available at http://doi.org/10.17605/OSF.IO/N4DKZ

### Authors’ contributions

**Samuel A Sutherland**: Conceptualisation, Investigation, Formal Analysis, Methodology, Software, Validation, Writing - original draft, Writing - Review & editing, Visualisation. **Jason Madan**: Supervision, Writing - review & editing. **Kat S Rock**: Methodology, Supervision, Writing - original draft, Writing - review & editing. All authors read and approved the final manuscript.

## Acknowledgements

Thanks to all of the HAT MEPP team and collaborators for the ongoing discussions which inspired the writing of this paper and for the development of the code that was used to generate the results presented here. The authors would like to thank Dr Erick Mwamba Miaka and the WHO HAT Atlas team for access to historical data from the DRC which allowed for the original model parameterisation in Antillon et al.[24]. The authors would also like to thank Dr Paul Bessell, Dr Vincent Jamonneau, Dr Veerle Lejon, Dr Elena Nicco, and Dr Iñaki Tirados for their input on the terminology used in this article.

For the purpose of open access, the authors have applied a Creative Commons Attribution (CC-BY) licence to any Author Accepted Manuscript version arising from this submission.

## Appendix A French-language version of this report

A French-language version of this article is available in the Open Science Framework repository as “`article_fr.pdf`”. This translation was created with the aid of DeepL, and was manually edited and verified by the authors.

## Appendix B Additional Results

All our results were generated under a range of eight different parameterisations. Equivalent figures to Figure 2 are available as a supplement. The code and raw data used to generate these figures are available in the Open Science Framework repository

## Appendix C Underlying model used for simulations

The model used is the same as the one used in Antillon et al. (2024) [24]. We used the ensemble parameterisations from this paper, modifying them slightly to remove the time-dependent improvement to passive surveillance and instead fixing it at the pre- or post-improvement level, depending on the strategy simulated. The following paragraph is a reproduction of the model description taken from the paper, for the convenience of the reader.

For this study, we used two variants of the previously published Warwick gHAT model [19, 26] consisting of a mechanistic, deterministic modelling framework to explicitly simulate transmission between humans and possibly animals via tsetse vectors (see the model formulation section of the supplement of Antillon et al.). Model parameterisation was performed individually for each model variant (models with and without possible animal transmission) and was updated compared to previous publications by fitting to WHO HAT Atlas data from 2000–2020 for each health zone in the DRC that had sufficient data—at least 10 or 13 data points for the models without and with animal transmission, respectively (where any year with AS and any year with non-zero passive case detection count as individual data points). This gave 165 health zones with a fit using the model without animal transmission and 156 that were fitted to both model variants. More details of the statistical fitting procedure are provided in the Supplementary Methods. The “ensemble” model consists of posteriors from both models. The proportion of samples from both models was determined by a statistical method (Bayes factors) that measures the relative goodness of fit of each model to the data.

