## Supplementary results for "Measuring elimination of *gambiense* human African trypanosomiasis: A comparison of deceptively different metrics"

Our simulations were run under eight different parameterisations, based on our existing modelling in the DRC. We use the model parameterisations from eight health zones, modified to have constant levels of passive surveillance at either the post-improvement value (for the strong PS strategies) or the pre-improvement value (for the weak PS strategies).

The figures in this supplement show various outputs from our model simulations and how they link to three different elimination metrics. Note that in the cases when not all realisations reach both WHO-EoT and LTE within the 200-year horizon of the simulation, the histogram only includes the realisations that successfully reach both metrics in order to calculate the difference. This means that the histograms should be taken with a level of caution when a large proportion of realisations do not result in elimination being reached by both of these metrics. Notably:

- Under the parameterisation for Yasa Bonga health zone (Figure 8), the third intervention strategy of weak passive screening with low levels of AS proved to be very slow to reach elimination by any of the metrics, and in the 200 years we simulated, only around 20% of realisations reached a state that could be called elimination.
- Under the parameterisation for Kwamouth health zone (Figure 5), the third intervention strategy of weak passive screening with low levels of AS proved to be insufficient to control the spread of $g$HAT within the 200-year horizon of our simulations, so only two of the 20 000 realisations ended up reaching both LTE and WHO-EoT.

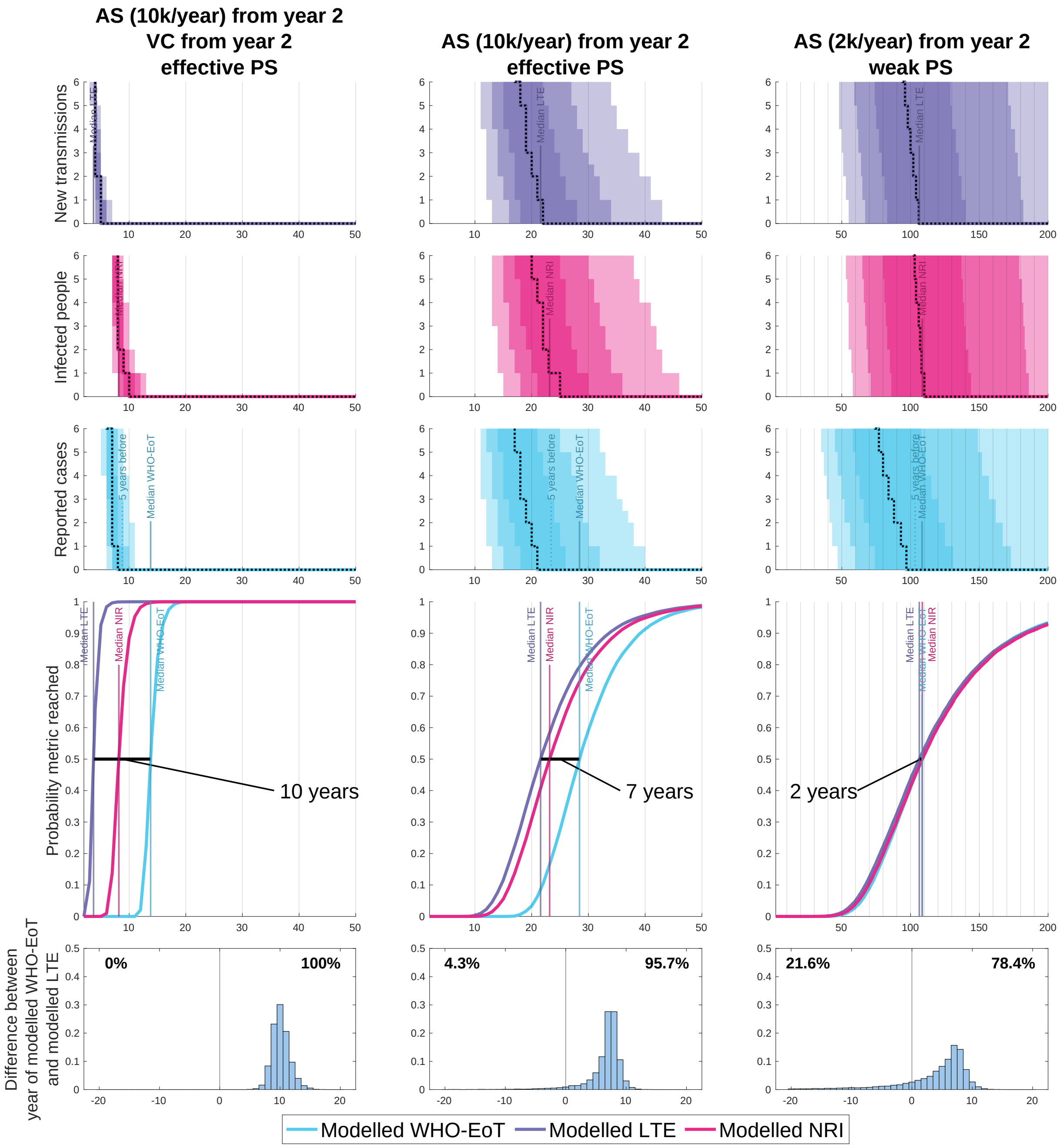


Figure 1: Model simulations showing various outputs and how they link to three different elimination metrics. The top three figures in each column show the number of new infections (purple) and the number of cases each year (blue), as well as the number of infections remaining at any given time (pink). The solid lines show the median model output, with 50%, 80%, and 95% prediction intervals shown by the shaded regions. The fourth figure shows estimates of the probability of Modelled-WHO-EoT, Modelled-LTE and Modelled-NRI by year, which are computed from the proportion of the 20 000 stochastic model simulations which have achieved the specific metric. The bottom figure shows a histogram of the delay between Modelled-LTE and WHO-EoT. The percentages show the proportion of realisations in which the final transmission event comes respectively after or before the five years of no cases required for WHO-EoT. The parameters used to generate the simulations used in this figure were based on our model parameterisation for **Bagata health zone in Bandundu Nord coordination** in the Democratic Republic of the Congo (DRC), although since we have not directly used case trends or intervention history here, this analysis does not reflect progress or predictions for the real health zone. [This Figure is the same as one main text figure, added here for easier comparison to the other parameterisations.]

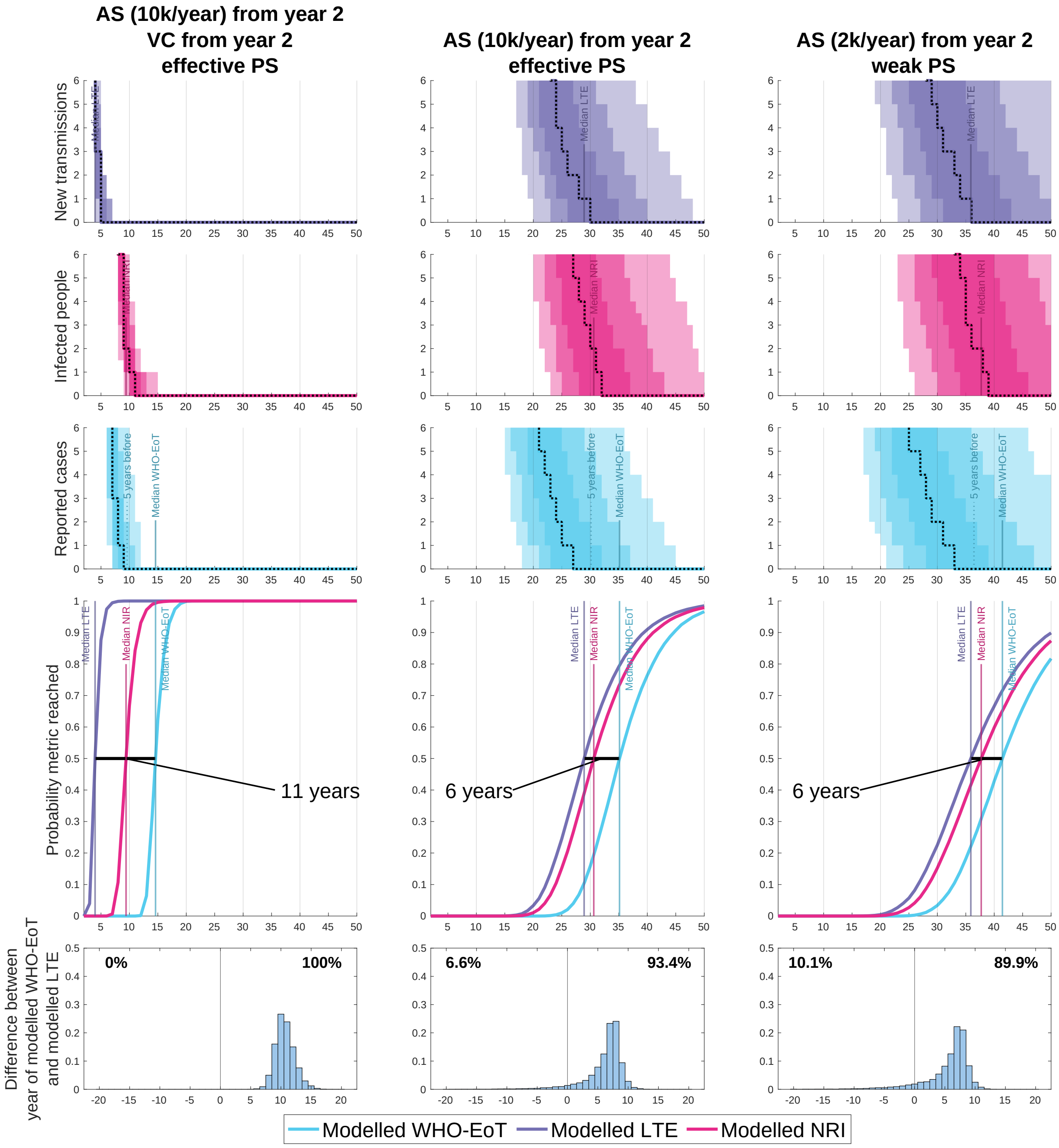


Figure 2: Model simulations showing various outputs and how they link to three different elimination metrics. The top three figures in each column show the number of new infections (purple) and the number of cases each year (blue), as well as the number of infections remaining at any given time (pink). The solid lines show the median model output, with 50%, 80%, and 95% prediction intervals shown by the shaded regions. The fourth figure shows estimates of the probability of Modelled-WHO-EoT, Modelled-LTE and Modelled-NRI by year, which are computed from the proportion of the 20 000 stochastic model simulations which have achieved the specific metric. The bottom figure shows a histogram of the delay between Modelled-LTE and WHO-EoT. The percentages show the proportion of realisations in which the final transmission event comes respectively after or before the five years of no cases required for WHO-EoT. The parameters used to generate the simulations used in this figure were based on our model parameterisation for **Boma Bungu health zone in Kongo Central coordination** in the DRC, although since we have not directly used case trends or intervention history here, this analysis does not reflect progress or predictions for the real health zone.

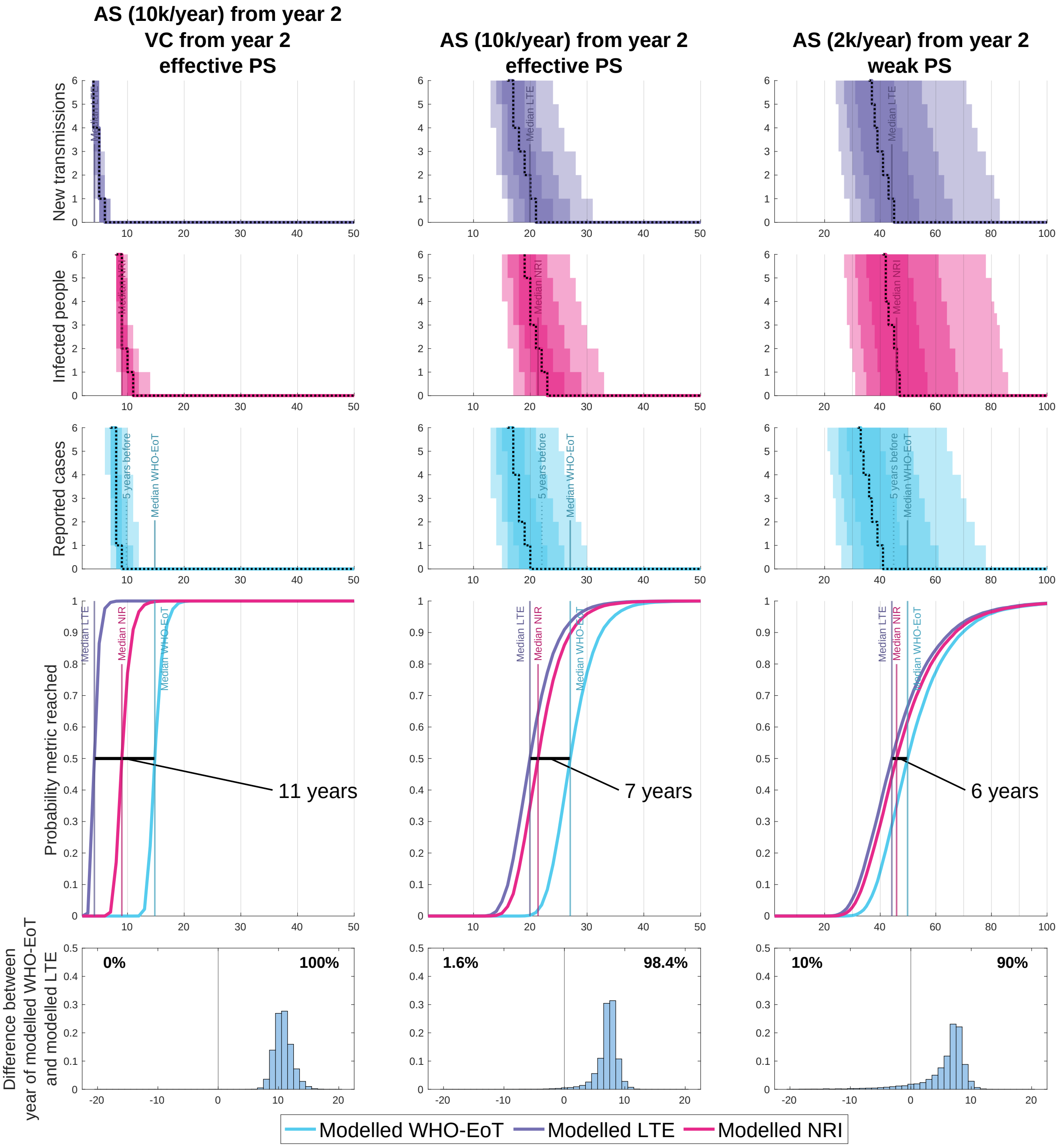


Figure 3: Model simulations showing various outputs and how they link to three different elimination metrics. The top three figures in each column show the number of new infections (purple) and the number of cases each year (blue), as well as the number of infections remaining at any given time (pink). The solid lines show the median model output, with 50%, 80%, and 95% prediction intervals shown by the shaded regions. The fourth figure shows estimates of the probability of Modelled-WHO-EoT, Modelled-LTE and Modelled-NRI by year, which are computed from the proportion of the 20 000 stochastic model simulations which have achieved the specific metric. The bottom figure shows a histogram of the delay between Modelled-LTE and WHO-EoT. The percentages show the proportion of realisations in which the final transmission event comes respectively after or before the five years of no cases required for WHO-EoT. The parameters used to generate the simulations used in this figure were based on our model parameterisation for **Bominenge health zone in Equateur Nord coordination** in the DRC, although since we have not directly used case trends or intervention history here, this analysis does not reflect progress or predictions for the real health zone.

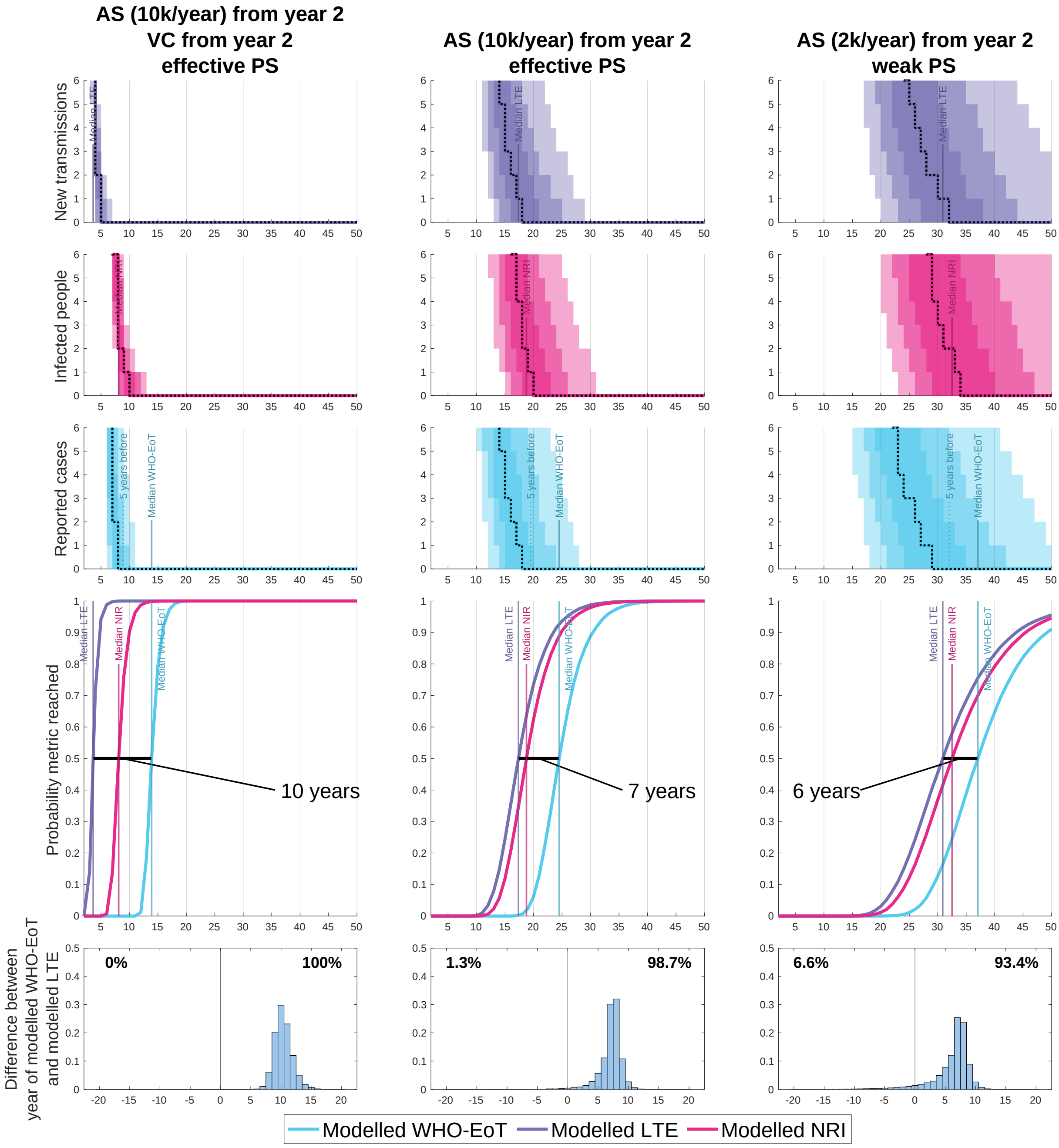


Figure 4: Model simulations showing various outputs and how they link to three different elimination metrics. The top three figures in each column show the number of new infections (purple) and the number of cases each year (blue), as well as the number of infections remaining at any given time (pink). The solid lines show the median model output, with 50%, 80%, and 95% prediction intervals shown by the shaded regions. The fourth figure shows estimates of the probability of Modelled-WHO-EoT, Modelled-LTE and Modelled-NRI by year, which are computed from the proportion of the 20 000 stochastic model simulations which have achieved the specific metric. The bottom figure shows a histogram of the delay between Modelled-LTE and WHO-EoT. The percentages show the proportion of realisations in which the final transmission event comes respectively after or before the five years of no cases required for WHO-EoT. The parameters used to generate the simulations used in this figure were based on our model parameterisation for **Budjala health zone in Equateur Nord coordination** in the DRC, although since we have not directly used case trends or intervention history here, this analysis does not reflect progress or predictions for the real health zone.

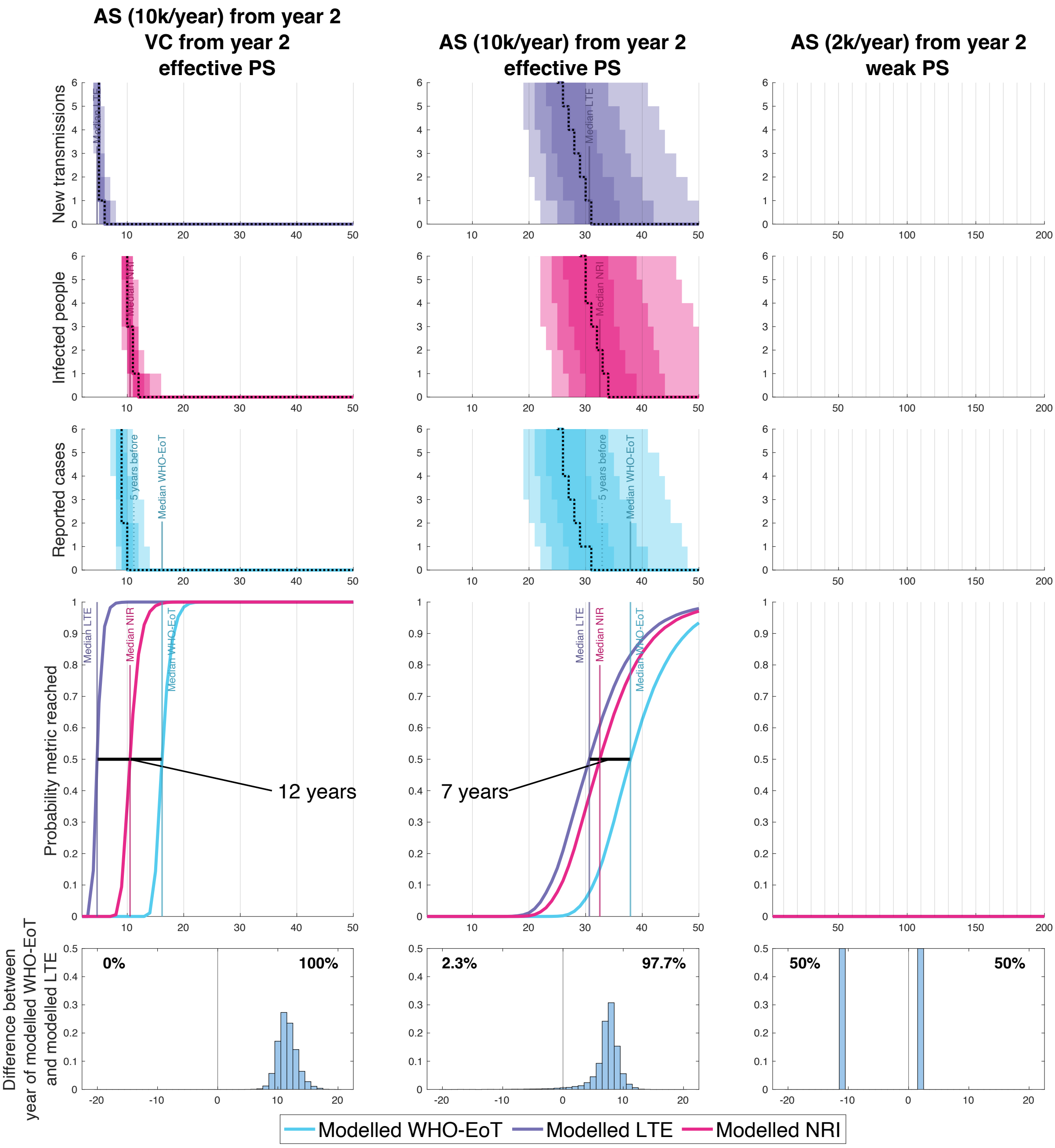


Figure 5: Model simulations showing various outputs and how they link to three different elimination metrics. The top three figures in each column show the number of new infections (purple) and the number of cases each year (blue), as well as the number of infections remaining at any given time (pink). The solid lines show the median model output, with 50%, 80%, and 95% prediction intervals shown by the shaded regions. The fourth figure shows estimates of the probability of Modelled-WHO-EoT, Modelled-LTE and Modelled-NRI by year, which are computed from the proportion of the 20 000 stochastic model simulations which have achieved the specific metric. The bottom figure shows a histogram of the delay between Modelled-LTE and WHO-EoT. The percentages show the proportion of realisations in which the final transmission event comes respectively after or before the five years of no cases required for WHO-EoT. The parameters used to generate the simulations used in this figure were based on our model parameterisation for **Kwamouth health zone in Bandundu Nord coordination** in the DRC, although since we have not directly used case trends or intervention history here, this analysis does not reflect progress or predictions for the real health zone.

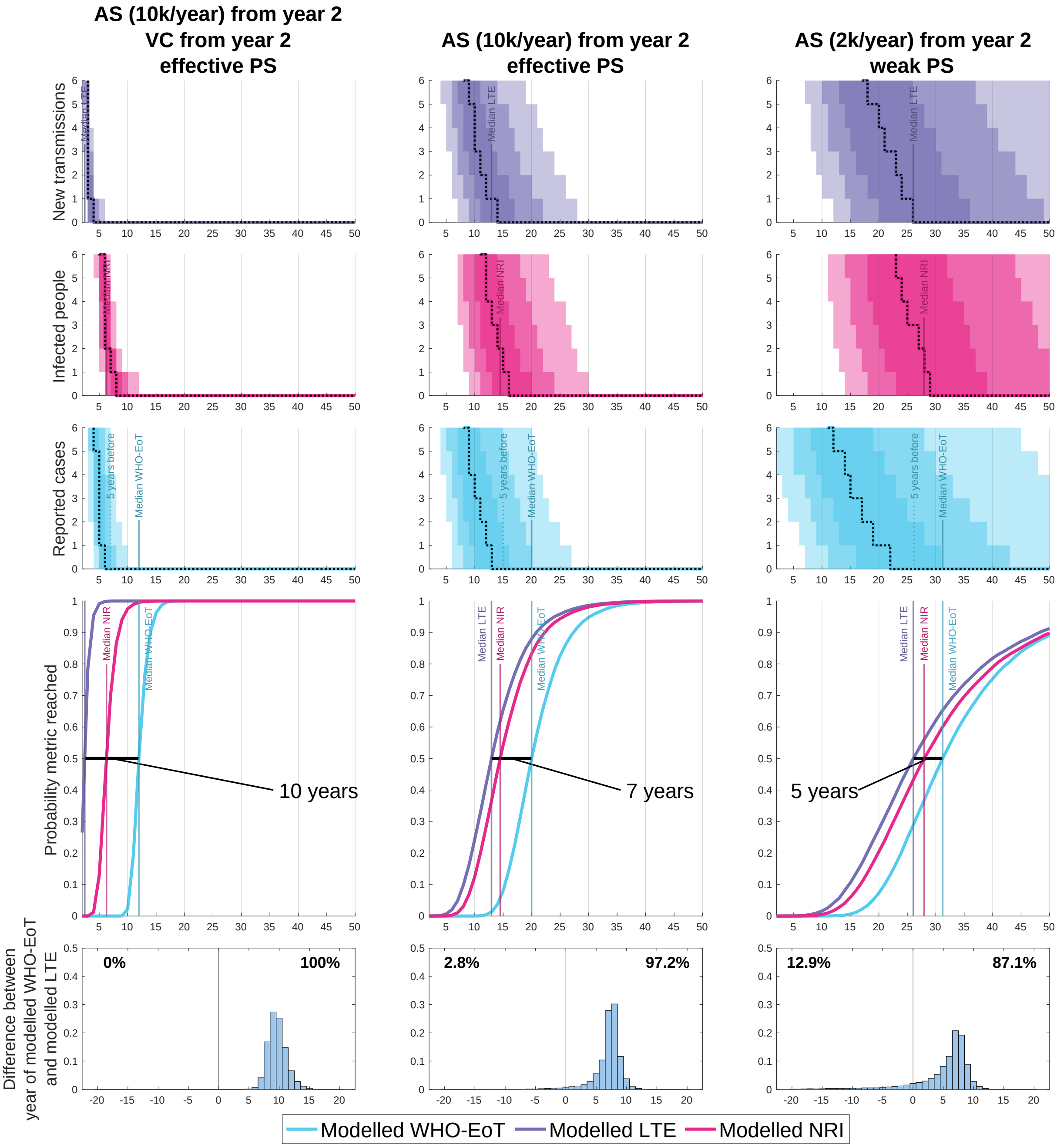


Figure 6: Model simulations showing various outputs and how they link to three different elimination metrics. The top three figures in each column show the number of new infections (purple) and the number of cases each year (blue), as well as the number of infections remaining at any given time (pink). The solid lines show the median model output, with 50%, 80%, and 95% prediction intervals shown by the shaded regions. The fourth figure shows estimates of the probability of Modelled-WHO-EoT, Modelled-LTE and Modelled-NRI by year, which are computed from the proportion of the 20 000 stochastic model simulations which have achieved the specific metric. The bottom figure shows a histogram of the delay between Modelled-LTE and WHO-EoT. The percentages show the proportion of realisations in which the final transmission event comes respectively after or before the five years of no cases required for WHO-EoT. The parameters used to generate the simulations used in this figure were based on our model parameterisation for **Mbaya health zone in Equateur Nord coordination** in the DRC, although since we have not directly used case trends or intervention history here, this analysis does not reflect progress or predictions for the real health zone.

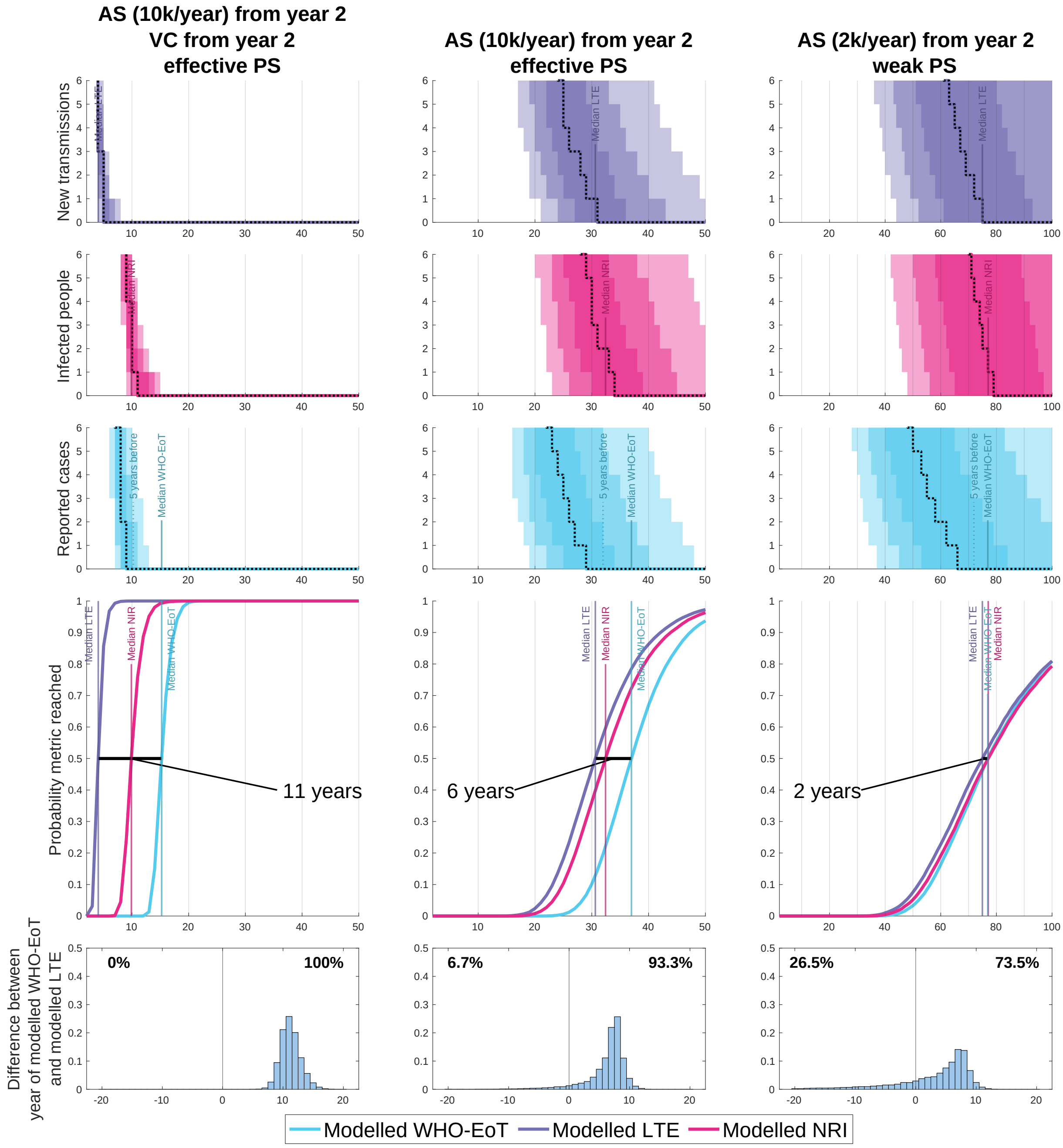


Figure 7: Model simulations showing various outputs and how they link to three different elimination metrics. The top three figures in each column show the number of new infections (purple) and the number of cases each year (blue), as well as the number of infections remaining at any given time (pink). The solid lines show the median model output, with 50%, 80%, and 95% prediction intervals shown by the shaded regions. The fourth figure shows estimates of the probability of Modelled-WHO-EoT, Modelled-LTE and Modelled-NRI by year, which are computed from the proportion of the 20 000 stochastic model simulations which have achieved the specific metric. The bottom figure shows a histogram of the delay between Modelled-LTE and WHO-EoT. The percentages show the proportion of realisations in which the final transmission event comes respectively after or before the five years of no cases required for WHO-EoT. The parameters used to generate the simulations used in this figure were based on our model parameterisation for **Mosango health zone in Bandundu Sud coordination** in the DRC, although since we have not directly used case trends or intervention history here, this analysis does not reflect progress or predictions for the real health zone.

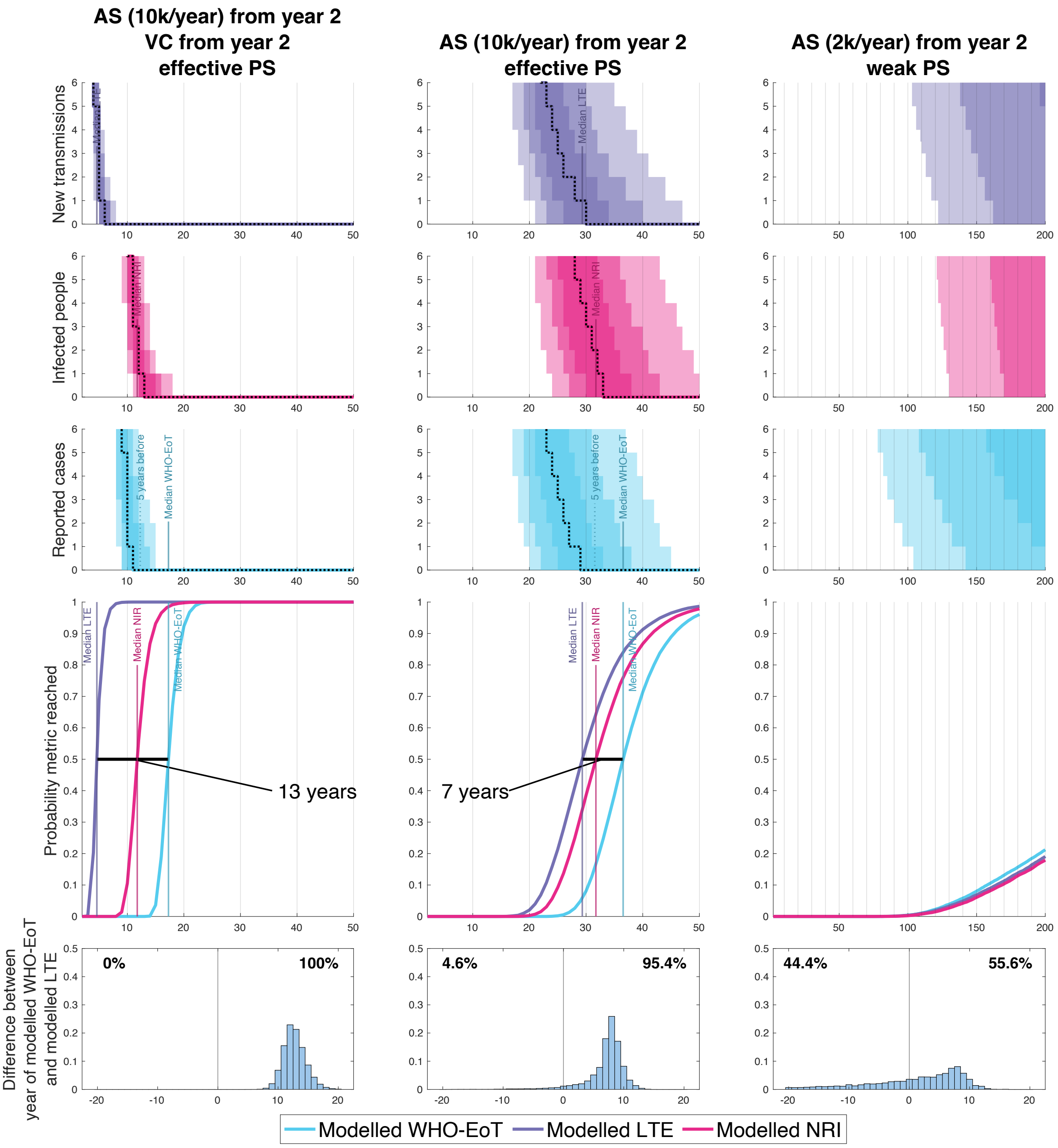


Figure 8: Model simulations showing various outputs and how they link to three different elimination metrics. The top three figures in each column show the number of new infections (purple) and the number of cases each year (blue), as well as the number of infections remaining at any given time (pink). The solid lines show the median model output, with 50%, 80%, and 95% prediction intervals shown by the shaded regions. The fourth figure shows estimates of the probability of Modelled-WHO-EoT, Modelled-LTE and Modelled-NRI by year, which are computed from the proportion of the 20 000 stochastic model simulations which have achieved the specific metric. The bottom figure shows a histogram of the delay between Modelled-LTE and WHO-EoT. The percentages show the proportion of realisations in which the final transmission event comes respectively after or before the five years of no cases required for WHO-EoT. The parameters used to generate the simulations used in this figure were based on our model parameterisation for **Yasa Bonga health zone in Bandundu Sud coordination** in the DRC, although since we have not directly used case trends or intervention history here, this analysis does not reflect progress or predictions for the real health zone.
